# A Retrospective Cohort Study of COVID-19 among Children in Fulton County, Georgia, March 2020 - June 2021

**DOI:** 10.1101/2021.10.30.21265680

**Authors:** Chloe M. Barrera, Mallory Hazell, Allison T. Chamberlain, Neel R. Gandhi, Udodirim Onwubiko, Carol Y. Liu, Juliana Prieto, Fazle Khan, Sarita Shah

**Author notes:** **Corresponding Author:** Chloe M. Barrera, MPH, 1518 Clifton Road, Atlanta, GA 30322, Emory University, Rollins School of Public Health, Department of Epidemiology. **Funding:** No outside funding sources were used for this article.

## Abstract

**Objective:** To describe case rates, testing rates, and percent positivity of coronavirus disease 2019 (COVID-19) among children aged 0-18 years by school-age grouping.

**Design:** We abstracted data from Georgia’s State Electronic Notifiable Disease Surveillance System on all 10,437 laboratory-confirmed COVID-19 cases among children aged 0-18 years during March 30, 2020 to June 6, 2021. We examined case rates, testing rates, and percent positivity by school-aged groupings, namely: preschool (0-4 years), elementary school (5-10 years), middle school (11-13 years), and high school (14-18 years) and compared these data among school aged children to those in the adult population (19 years and older).

**Setting:** Fulton County, Georgia.

**Main outcome measures:** COVID-19 case rates, testing rates, and percent positivity.

**Results:** Over time, the proportion of pediatric cases rose substantially from 1.1% (April 2020) to 21.6% (April 2021) of all cases in the county. Age-specific case rates and test rates were consistently highest among high-school aged children. Test positivity was similar across school-age groups, with periods of higher positivity among high-school aged children.

**Conclusions:** Low COVID-19 testing rates among children, especially early in the pandemic, likely underestimate the true burden of disease in this age group. Despite children having lower measured incidence of COVID-19, we found when broader community incidence increased, incidence also increased among all pediatric age groups. As the COVID-19 pandemic continues to evolve, it remains critical to continue learning about the incidence and transmissibility of COVID-19 in children.

## Introduction

In the United States, fewer cases of the coronavirus disease 2019 (COVID-19) have been reported in children as compared to adults.^1^ Prior to the surge of the Delta variant, children accounted for approximately 19% of all cases.^1^ While early evidence found children to have lower susceptibility to COVID-19,^2^ as well as milder cases and overall better prognosis than adults,^3–5^ children still experienced severe outcomes. For example, as of March 2021, 2,617 cases of COVID-19 associated multisystem inflammatory syndrome in children had been reported in the United States, of which 1.3% resulted in death^6^ and 2.1% of all pediatric COVID-19 cases resulted in hospitalization and 0.03% resulted in death.^7^ Further, it had been reported one in three children who were hospitalized for COVID-19 had a severe case resulting in admission to a pediatric intensive care unit.^8,9^

Early pediatric data were often reported in aggregate by age, limiting our ability to learn about viral transmission by more granular pediatric age groups with distinct COVID-19 epidemiology. Aggregation of pediatric data obscures important epidemiologic associations and trends useful to constituents like childcare providers and school adminstrators.^10^ Specifically, disaggregating data by age becomes particularly important for determination of the opening and closing of schools and classrooms given younger children are more likely to be asymptomatic and have lower rates of onward transmission^11^ while older children transmit the virus at rates more similar to adults.^12^ Finally, early in the pandemic, many testing venues chose not to test toddler or preschool aged children,^13^ and there was concern selective testing among all pediatric age groups would lead to lower measured rates of COVID-19 in children than adults.

Fulton County, the largest county in the state of Georgia, is a core county of metropolitan Atlanta, and includes 90% of the City of Atlanta as well as several suburban cities (estimated population 1.1 million).^14^ Over the course of the pandemic, periods of lower and higher COVID-19 cases in Fulton County have mirrored those of the state of Georgia, though Fulton County has had a lower rate of COVID-19 cases per capita.^15^ The county reported its first COVID-19 case on March 2, 2020, and has since offered free COVID-19 testing to eligible residents. In accordance with the Centers for Disease Control and Prevention and Georgia Department of Public Health (GDPH) guidelines, testing was initially restricted to symptomatic persons or those with risk factors for severe disease, but included children of all ages. Eligibility was expanded in April 2020 to include high-risk workers regardless of symptoms. By May 2020, eligibility for testing expanded to include anyone who desired testing, regardless of age, risk group, or symptoms. The wide availability of free and accessible COVID-19 testing in Fulton County provides a unique opportunity to understand the epidemiology of COVID-19, with reduced artifact from restrictive testing policies in other public health jurisdictions and states. We describe trends in rates of COVID-19 cases, testing, and test positivity, prior to the Delta surge, among children in Fulton County from March 2020 – June 2021.

## Methods

### Data Source and Study Sample

This retrospective cohort study used data from children aged 0-18 years and adults aged 19 years and older diagnosed with laboratory-confirmed SARS-CoV-2 Infection (the coronavirus that causes COVID-19) from March 30, 2020 to June 6, 2021, who resided in Fulton County. A laboratory confirmation for SARS-CoV-2 was defined as a positive result of real-time reverse transcriptase–polymerase chain reaction test.

Data were extracted from the State Electronic Notifiable Disease Surveillance System (SendSS) on September 24, 2021 to allow sufficient time for case investigations to be completed. SendSS is an electronic database developed by GDPH to track patients with notifiable diseases, including case investigation of COVID-19 patients across the state. Through the case investigation process, cases are asked a series of standardized questions about their COVID-19 experience, including questions about close contacts.^16^ Any demographic information not provided through the electronic lab result is also collected. For cases under 18, interviews are conducted with a parent, guardian, or with the case themselves if parental approval was obtained.

We extracted the following data for each case: date of first SARS-CoV-2 positive specimen collection, age, gender, race, ethnicity, hospitalization, intensive care unit admission, death, symptoms, and potential exposure.

Data from Georgia’s Online Analytical Statistical Information System^14^ were used to obtain the number of children and adults residing in Fulton County. These data were used to determine age-specific per capita rates of COVID-19 cases and COVID-19 tests administered by age group; children were grouped by ages 0-4, 5-10, 11-13, and 14-18 years and adults were grouped by ages 19 years and older. The age stratification for children corresponds to school-age groupings allowing us to draw inferences about differences in rates of COVID-19 based on school-age.

### Statistical Analysis

We used descriptive statistics to report differences in demographic characteristics, COVID-19 outcomes, symptoms, and exposures by school-age group. We examined the proportion of cases attributable to children by 14-day periods among all children. We also examined age-specific per capita case rates and test rates (i.e. number of cases and number of tests per 10,000 individuals) and testing percent positivity by 14-day periods among children by school-age group and among adults aged 19 years and older.

### Patient and Public Involvement Statement

Patients and/or the public were not involved in the design, or conduct, or reporting, or dissemination plans of this research.

### Ethical considerations

As a public health surveillance activity in response to the COVID-19 emergency response, this activity was deemed exempt from IRB by the GDPH Review Board.

## Results

From March 30, 2020 to June 6, 2021, there were 10,437 pediatric cases of COVID-19 reported in Fulton County, a case rate of 431.4 cases per 10,000 children aged 0-18 years (over this same time period there were 73,543 adult cases of COVID-19 reported in Fulton County, a case rate of 880.3 cases per 10,000 adults aged 19 years and older). Among children, almost half of the cases were aged 14-18 years (48.3%), 16.9% were 11-13 years, 21.7% were 5-10 years, and 13.1% were 0-4 years (Table 1). Across pediatric age groups, there were similar numbers of female (50.2%) and male cases (49.4%). The most common race/ethnicities across pediatric age groups were non-Hispanic Black (35.0%) and non-Hispanic White (33.5%).

**Table 1.** Demographic and Clinical Characteristics of Reported COVID-19 Cases among Children Aged 0-18 Years in Fulton County, GA from March 30, 2020 - June 6, 2021

| **Characteristics** | **All Cases** | **Cases by Pediatric Age Group** |  |  |  |
| --- | --- | --- | --- | --- | --- |
|  | 0-18 | 0-4 | 5-10 | 11-13 | 14-18 |
| **Total, n (%)** | 10,437 (100.0) | 1367 (13.1) | 2261 (21.7) | 1767 (16.9) | 5042 (48.3) |
| **Gender** |  |  |  |  |  |
| Female | 5238 (50.2) | 670 (49.0) | 1117 (49.4) | 884 (50.0) | 2567 (50.9) |
| Male | 5160 (49.4) | 689 (50.4) | 1134 (50.2) | 880 (49.8) | 2457 (48.7) |
| Unknown | 39 (0.4) | 8 (0.6) | 10 (0.4) | 3 (0.2) | 18 (0.4) |
| **Race/Ethnicity** |  |  |  |  |  |
| Asian, non-Hispanic | 419 (4.0) | 54 (4.0) | 89 (3.9) | 68 (3.8) | 208 (4.1) |
| Black, non-Hispanic | 3655 (35.0) | 555 (40.6) | 892 (39.5) | 618 (35.0) | 1590 (31.5) |
| Hispanic, all races | 1442 (13.8) | 193 (14.1) | 346 (15.3) | 293 (16.6) | 610 (12.1) |
| White, non-Hispanic | 3494 (33.5) | 348 (25.5) | 577 (25.5) | 547 (31.0) | 2022 (40.1) |
| Other, non-Hispanic | 496 (4.8) | 75 (5.5) | 118 (5.2) | 92 (5.2) | 211 (4.2) |
| Unknown | 931 (8.9) | 142 (10.4) | 239 (10.6) | 149 (8.4) | 401 (8.0) |
| **Hospitalized** |  |  |  |  |  |
| Yes | 132 (1.3) | 49 (3.6) | 23 (1.0) | 6 (0.3) | 54 (1.1) |
| No | 10305 (98.7) | 1318 (96.4) | 2238 (99.0) | 1761 (99.7) | 4988 (98.9) |
| **Intubated** |  |  |  |  |  |
| Yes | 18 (0.2) | 4 (0.3) | 4 (0.2) | 0 (0.0) | 10 (0.2) |
| No | 10419 (99.8) | 1363 (99.7) | 2257 (99.8) | 1767 (100.0) | 5032 (99.8) |
| **Died** |  |  |  |  |  |
| Yes | 1 (<0.0) | 0 (0.0) | 0 (0.0) | 0 (0.0) | 1 (<0.0) |
| No | 10,436 (99.9) | 1367 (100.0) | 2261 (100.0) | 1767 (100.0) | 2485 (99.9) |

Hospitalizations were most common among children aged 0-4 years, occurring in 3.6% of cases. Across all pediatric age groups, less than 0.5% of cases were intubated and one case (aged 14-18 years) died.

Case investigation or medical record review was completed for 7484 (71.7%) pediatric cases, providing additional information on symptoms and potential exposures (Table 2). Less than half of all children reported COVID-19 symptoms at diagnosis (43.2%). The most commonly reported symptoms across all ages were cough (17.2%) and fever (16.1%); headaches were commonly reported among older children (25.4% among children aged 14-18 years but not among younger children (3.8% among children aged 0-4 years).

**Table 2.** Symptomaticity and Exposure Mechanisms of Reported COVID-19 Cases among Children Aged 0-18 Years with Completed Case Investigations in Fulton County, GA from March 30, 2020 - June 6, 2021

| **Characteristics** | **All Cases** | **Cases by Pediatric Age Group** |  |  |  |
| --- | --- | --- | --- | --- | --- |
|  | 0-18 | 0-4 | 5-10 | 11-13 | 14-18 |
| **Total, n (%)<sup>a</sup>** | 7484 (100.0) | 1030 (13.8) | 1699 (22.7) | 1286 (17.2) | 3469 (46.3) |
| **Symptomaticity** |  |  |  |  |  |
| Symptomatic | 4507 (43.2) | 593 (43.4) | 851 (37.6) | 781 (44.2) | 2282 (45.3) |
| Asymptomatic | 5930 (56.8) | 774 (56.6) | 1410 (62.4) | 986 (55.8) | 2760 (54.7) |
| **Cough** |  |  |  |  |  |
| Yes | 1794 (17.2) | 289 (21.1) | 288 (12.7) | 278 (15.7) | 939 (18.6) |
| No | 4903 (47.0) | 620 (45.4) | 1218 (53.9) | 913 (51.7) | 2152 (42.7) |
| **Shortness of breath/difficulty breathing** |  |  |  |  |  |
| Yes | 403 (3.9) | 42 (3.1) | 40 (1.8) | 43 (2.4) | 278 (5.5) |
| No | 6210 (59.5) | 832 (60.9) | 1457 (64.4) | 1144 (64.7) | 2777 (55.1) |
| **Fever** |  |  |  |  |  |
| Yes | 1677 (16.1) | 327 (23.9) | 371 (16.4) | 267 (15.1) | 712 (14.1) |
| No | 5023 (48.1) | 590 (43.2) | 1143 (50.6) | 928 (52.5) | 2362 (46.8) |
| **Chills** |  |  |  |  |  |
| Yes | 788 (7.6) | 45 (3.3) | 98 (4.3) | 125 (7.1) | 520 (10.3) |
| No | 5789 (55.5) | 814 (59.5) | 1391 (61.5) | 1052 (59.5) | 2532 (50.2) |
| **Muscle pain** |  |  |  |  |  |
| Yes | 1149 (11.0) | 40 (2.9) | 148 (6.5) | 183 (10.4) | 778 (15.4) |
| No | 5439 (52.1) | 810 (59.3) | 1349 (59.7) | 994 (56.3) | 2286 (45.3) |
| **Headache** |  |  |  |  |  |
| Yes | 2108 (20.2) | 44 (3.2) | 384 (17.0) | 399 (22.6) | 1281 (25.4) |
| No | 4509 (43.2) | 802 (58.7) | 1120 (49.5) | 786 (44.5) | 1801 (35.7) |
| **Sore throat** |  |  |  |  |  |
| Yes | 1460 (14.0) | 54 (4.0) | 214 (9.5) | 254 (14.4) | 938 (18.6) |
| No | 5159 (49.4) | 806 (59.0) | 1284 (56.8) | 937 (53.0) | 2132 (42.3) |
| **Abdominal pain** |  |  |  |  |  |
| Yes | 358 (3.4) | 42 (3.1) | 80 (3.5) | 69 (3.9) | 167 (3.3) |
| No | 5643 (54.1) | 756 (55.3) | 1303 (57.6) | 1008 (57.0) | 2576 (51.1) |
| **Nausea or vomiting** |  |  |  |  |  |
| Yes | 580 (5.6) | 77 (5.6) | 113 (5.0) | 97 (5.5) | 293 (5.8) |
| No | 6049 (58.0) | 811 (59.3) | 1388 (61.4) | 1089 (61.6) | 2761 (54.8) |
| **Diarrhea** |  |  |  |  |  |
| Yes | 437 (4.2) | 106 (7.8) | 74 (3.3) | 63 (3.6) | 194 (3.8) |
| No | 6189 (59.3) | 786 (57.5) | 1420 (62.8) | 1123 (63.6) | 2860 (56.7) |
| **Other** |  |  |  |  |  |
| Yes | 751 (7.2) | 95 (6.9) | 100 (4.4) | 109 (6.2) | 447 (8.9) |
| No | 4690 (44.9) | 637 (46.6) | 1142 (50.5) | 881 (49.9) | 2030 (40.3) |
| **Exposure through a household contact** |  |  |  |  |  |
| Yes | 2859 (27.4) | 483 (35.3) | 862 (38.1) | 544 (30.8) | 970 (19.2) |
| No | 2994 (28.7) | 331 (24.2) | 536 (23.7) | 497 (28.1) | 1630 (32.3) |
| **Exposure through a community contact** |  |  |  |  |  |
| Yes | 1406 (13.5) | 176 (12.9) | 275 (12.2) | 230 (13.0) | 725 (14.4) |
| No | 3815 (36.6) | 554 (40.5) | 990 (43.8) | 704 (39.8) | 1567 (31.1) |

<sup>a</sup>Percentages do not add up to 100 due to unknown or missing data.

Exposure to COVID-19 through a household contact was reported more often for younger children (35.3% among children aged 0-4 years) than for older children (19.2% among children aged 14-18 years), whereas exposure to COVID-19 through a community contact was more similar across age groups (13.5%) (Table 2).

Over time, the proportion of pediatric cases rose substantially and surpassed the proportion of children aged 0-18 years in the population of Fulton County (9.2%).^14^ In early April 2020, children aged 0-18 years represented 1.1% of all cases in the county, by September 2020 they represented 15.4% of all cases in the county, and by the end of April 2021 they represented 21.6% of all cases in the county (Figure 1). The most recent data in this report showed a decline in cases among children aged 0-18 years; in early June 2021 they represented 11.8% of all cases in the county.

**Figure 1.**
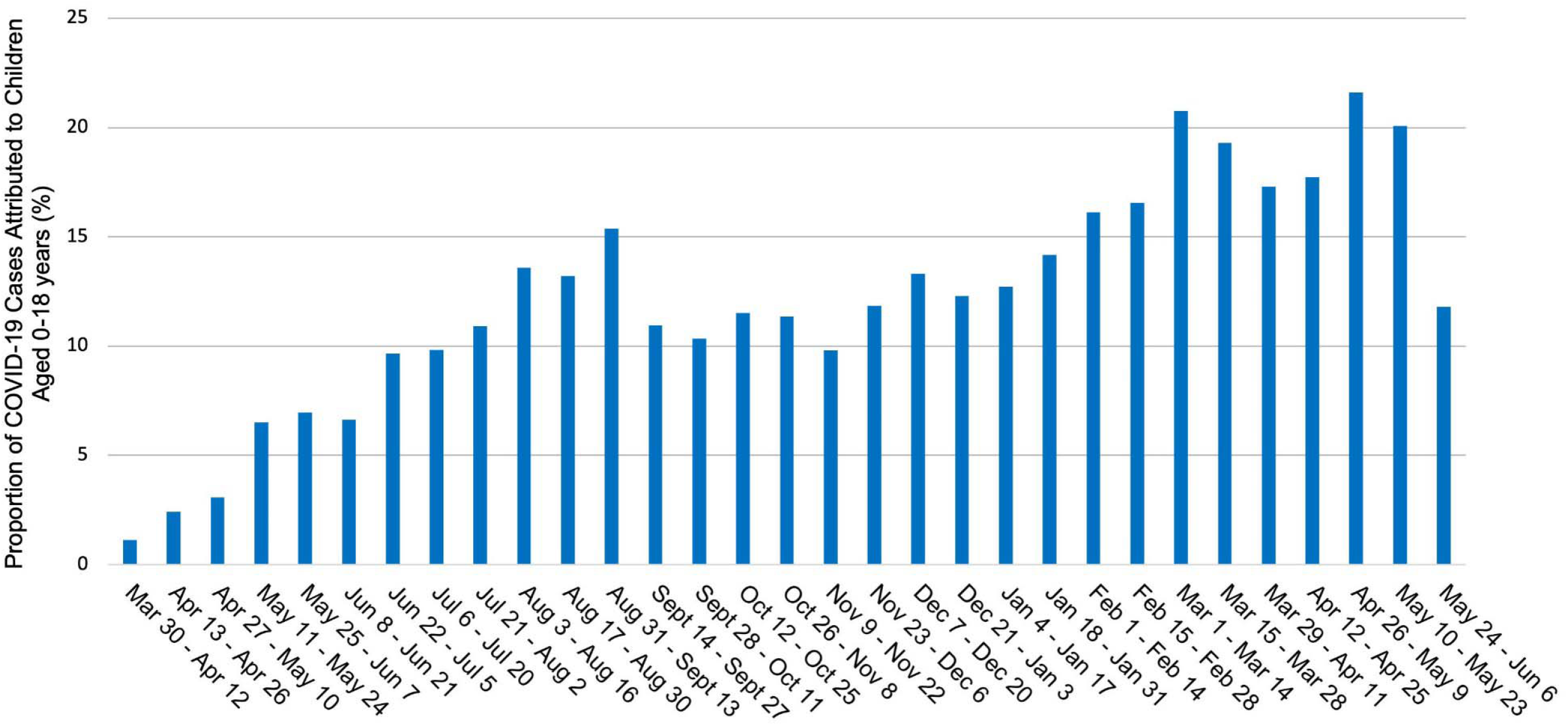
Proportion of COVID-19 Cases Attributed to Children Aged 0-18 Years in Fulton County, Georgia by 14-day Periods, March 2020 - June 2021

Age-specific case rates (Figure 2) were consistently highest among adults and children aged 14-18 years. There were periods of time when case rates among children aged 14-18 years were higher than among adults (August 2020 and March 2021). Testing rates were consistently highest among adults (Figure 3). Among children, testing rates were consistently highest among those aged 14-18 years compared to younger children. Data from December 7-20, 2020 show more than 2.5 times the number of tests given to children aged 14-18 years (554 tests per 10,000 children aged 14-18 years) compared to children aged 0-4 years (208 tests per 10,000 children aged 0-4 years). Despite these marked differences in testing rates, test positivity was similar across pediatric age groups and percent positivity among pediatric cases was higher than adults at almost every time period observed after July, 2020 (Figure 4).

**Figure 2.**
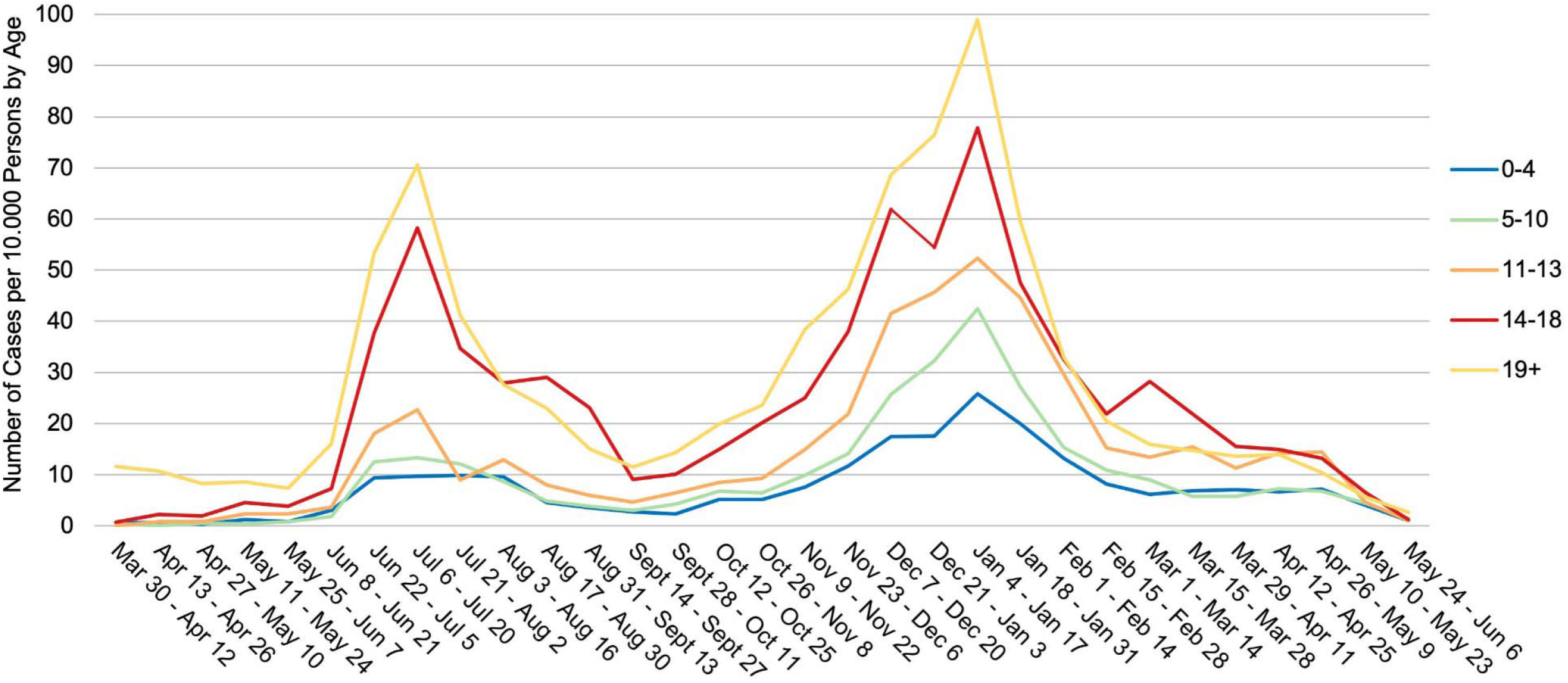
Age Distribution of COVID-19 Cases per 10,000 Persons by Age Group in Fulton County, Georgia by 14-day Periods, March 2020 - June 2021

**Figure 3.**
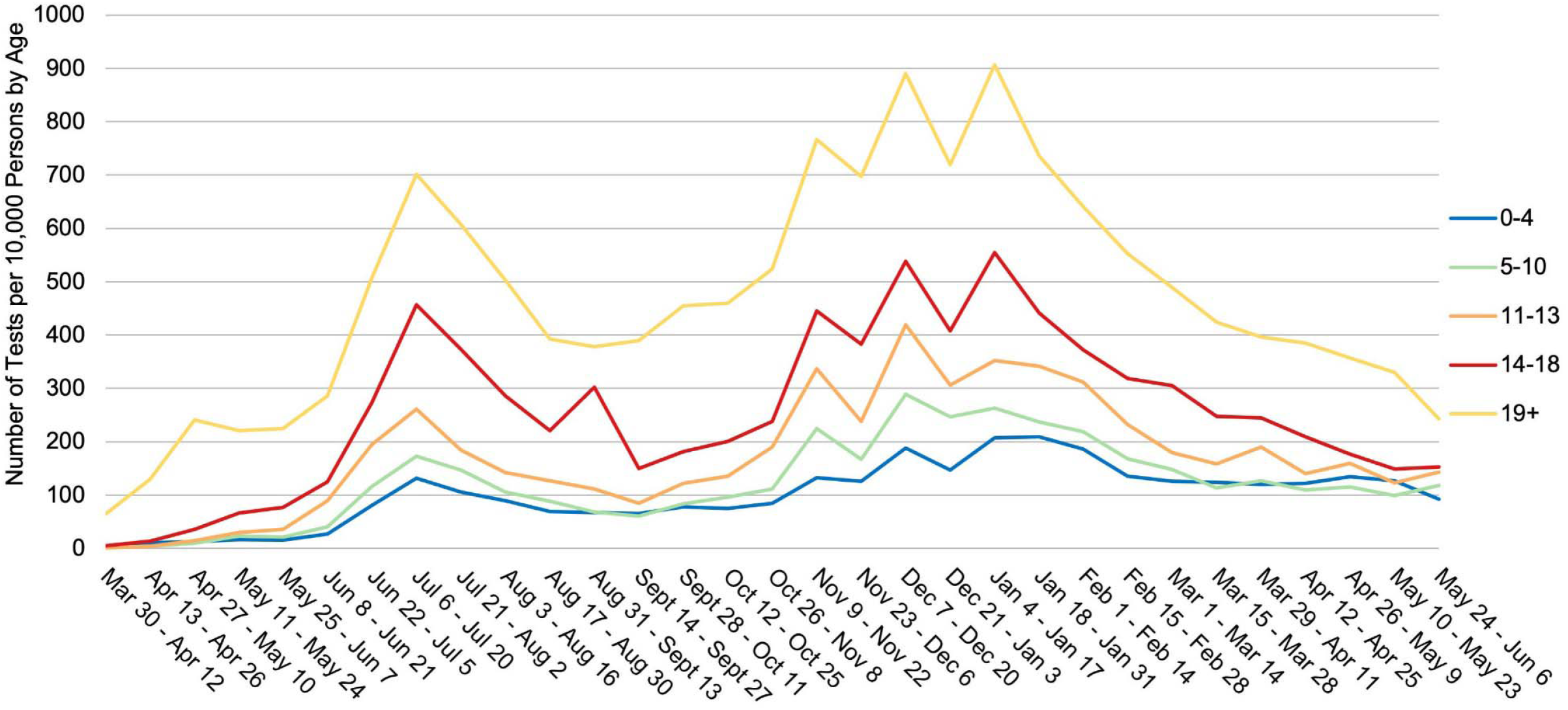
COVID-19 Tests per 10,000 Persons by Age Group in Fulton County, Georgia by 14-day Periods, March 2020 - June 2021

**Figure 4.**
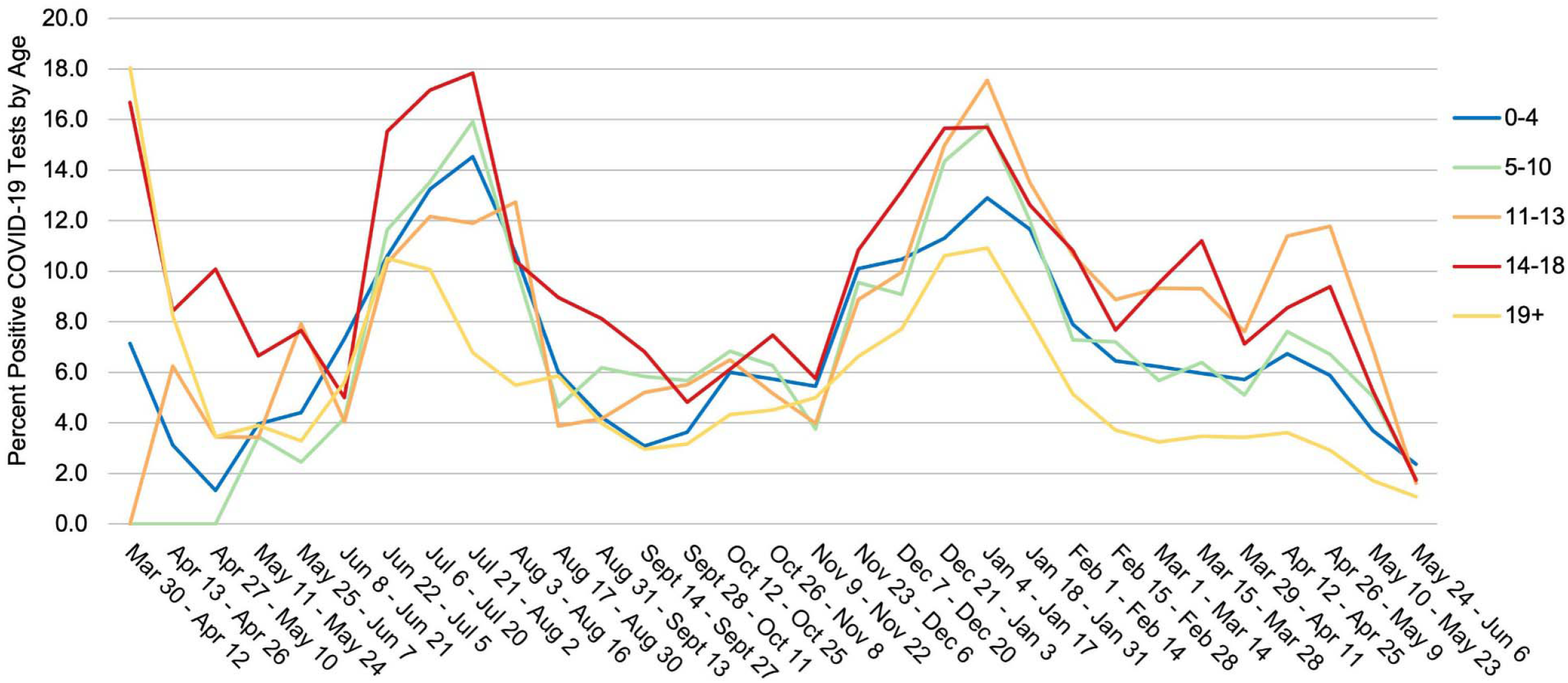
Percent Positive COVID-19 Tests by Age Group in Fulton County, Georgia by 14-day Periods, March 2020 - June 2021

## Discussion

This report contributes a comprehensive picture of pediatric COVID-19 by school-age groupings in Fulton County prior to the emergence of the Delta variant. While some published studies present data on case prevalence, we provide additional insights on testing rates and test positivity to give a more complete view of COVID-19 among children. When testing became more widely available for all ages regardless of symptoms in June 2020, we found the proportion of COVID-19 cases among all children remained at or above 10%. Given children in our population were tested less than half as frequently as adults, our pediatric case rates likely underestimated the true burden of COVID-19 among this age group. Our data on test positivity further underscore this hypothesis: among children, testing and case numbers were highest among older children; however, younger children had similar rates of COVID-19 test positivity as older children and all children had similar rates of COVID-19 test positivity as adults, suggesting less frequent testing may have underestimated the true prevalence of COVID-19 cases among children.

With case interviews completed for 72% of children diagnosed with COVID-19, we found a substantial proportion (56.8%) were asymptomatic at the time of their COVID-19 diagnosis. It has been estimated 79% of infections in 10-to 19-year-olds are asymptomatic;^11^ further suggesting our estimates on diagnosed case rates represented only the “tip of the iceberg” compared to true pediatric infections. In line with previous studies of the clinical manifestation of COVID-19 in this age group,^17^ fever and cough were commonly reported among all age groups with older children more commonly reporting headache, muscle pain, or sore throat. Importantly, clinical outcomes for children were overwhelmingly favorable, 1.3% of cases were hospitalized and one case died. Given the lower rates of testing among younger children and likely under-diagnosis of COVID-19 disease in this age group, it is highly likely that true pediatric infection hospitalization rate and infection fatality rate was even lower than what we observed. One way to obtain a better estimate of the true infection rate is to implement universal testing; while this was not implemented in Fulton County, estimated seroprevalence data from Georgia suggested cumulative infections rates of SARS-CoV-2 among those aged less than 18 years were similar to the general population (16.0% vs. 17.8%, respectively).^1^

Similar to previous studies,^18^ we found when broader community incidence increases, incidence among all age groups increases suggesting community spread impacts children as well as adults; however, it is important to recognize increases in testing are commensurate with increases in community prevalence, a phenomenon which suggests testing volume may be a marker for increased COVID-19 community spread, exposure, or risk-taking behavior at the population level. Understanding the potential role children play in the COVID-19 pandemic and carefully monitoring case rates, testing rates, and test positivity trends in children and the factors that drive transmissibility among children is critical.

Events over 2020 and the first half of 2021 can help explain points in time when we saw higher and lower rates of COVID-19 among children. All Fulton County schools closed on March 10, 2020 and did not reopen for the remainder of the academic year. The first large spike in pediatric cases, observed in late June to early-July 2020, may be partially attributed to summer travel or children attending summer camps, particularly among children aged 14-18 years. During June 17-20, 2020 one overnight camp in Georgia resulted in 260 known cases of COVID-19; 51% of those aged 6-10 years and 44% of those aged 11-17 years were infected.^19^ Test results were only available for 58% of attendees, so the true impact of this outbreak was likely underestimated, but this outbreak was highly publicized and may have resulted in an acute pre- and post-camp testing surge. Additionally, on July 8, 2020, the Mayor of Atlanta signed an executive order mandating the use of face masks in public spaces and also prohibited gatherings of more than ten people, factors known to reduce transmission of viruses.^20^ The potential impact of this order, together with other changes in travel patterns, may explain the downward trends observed from the end of July through September 2020. In September 2020, Fulton County schools began phasing-in in-person instruction in a hybrid format (i.e. both in-person and virtual instruction options) and by October 14, 2020 full-time in-person instruction was offered but remained optional. Pediatric cases increased again substantially from early November 2020 through mid-January 2021, mirroring the trends observed in the larger community. Factors contributing to this winter surge may have included the return of in-person schooling, holiday travel and mixing of households’ indoors, and potentially unrecognized transmission of novel variants such as B.1.1.7, which was first identified in the United States in December 2021.^21^ Since the 2021 winter surge, pediatric cases declined through the start of June 2021 (the latest date for which this paper contains data). Availability and uptake of COVID-19 vaccines, which were available to all individuals in Georgia over the age of 16 beginning March 25, 2021 and over the age of 12 beginning May 11, 2021 may help explain some of the downward trend in COVID-19 incidence the county experienced.

This evaluation was subject to limitations. First, while Fulton County Board of Health offered testing to individuals of all ages beginning in May, 2020, not all testing facilities in and around Fulton County made COVID-19 testing readily available to children. This is an important factor that likely contributed substantially to the number of pediatric tests being far fewer than the number of adult tests. Second, data available from case investigations of young children depend entirely on interviews with proxies, which do not always result in the most accurate data on symptomaticity or source of infection. It is also important to acknowledge we did not have data on whether children were attending school in-person during this time frame, limiting an ability to make inferences about the role schools may have played in pediatric COVID-19 transmission; however, studies have revealed low rates of COVID-19 transmission within schools when mitigation strategies are in place.^22,23^

Strengths of this study include our data sources. In Georgia, SendSS is the most comprehensive surveillance data source for COVID-19 cases and includes data on cases tested across the majority of testing sites in Fulton County. This allowed us to observe pediatrics trends using the most data possible for cases diagnosed among Fulton County residents, regardless of testing location. Because our datasets included case age, we could also examine COVID-19 trends among smaller, informative pediatric age groups.

## Conclusion

This in-depth epidemiologic study on children in Fulton County adds to the emerging data on COVID-19 in children. While it has been thought children are less susceptible to COVID-19 than adults, our data on testing rates and test positivity indicate the true burden of COVID-19 among those aged 18 years and younger may have been underestimated prior to the Delta variant. Our observations provide a platform for further detailed studies on the epidemiology of COVID-19 in children.

### What’s Known on This Subject

Prior to the Delta variant, studies consistently demonstrated children’s susceptibility to COVID-19; however, incidence and severe outcomes were lower among children than adults.

### What This Study Adds

Children tested positively at similar rates to adults indicating low testing rates likely underestimated the true burden of pediatric COVID-19 prior to the Delta variant.

## Data Availability

All data produced in the present study are protected given they are identifiable.

## Contributor Statements

Ms. Barrera conceptualized and designed the study, analyzed and interpreted the data, drafted the initial manuscript, and critically revised the manuscript.

Ms. Hazell and Ms. Onwubiko organized the data, carried out the initial analyses, and revised the manuscript.

Drs. Shah and Chamberlain conceptualized and designed the study and reviewed and revised the manuscript.

Drs. Gandhi and Khan and Ms. Prieto contributed subject matter expertise and reviewed and revised the manuscript.

All authors approved the final manuscript as submitted and agree to be accountable for all aspects of the work.

